# The role of comorbidities and clinical predictors of severe disease in COVID-19: a systematic review and meta-analysis

**DOI:** 10.1101/2020.04.21.20074633

**Authors:** Reza Tabrizi, Kamran B Lankarani, Peyman Nowrouzi-sohrabi, Mojtaba Shabani-Borujeni, Shahla Rezaei, Mahnaz Hosseini-bensenjan, Sina vakili, Seyed Taghi Heydari, Mohammad Ali Ashraf

## Abstract

**Background:** COVID_19 is unpredictable due to non-specific symptoms and clinical course diversity in different individuals. We analyzed studies regarding the factors associated with severe status of the disease to identify unique findings in severely affected patients.

**Methods:** We systematically searched the electronic databases, including PubMed, Scopus, EMBASE, Web of Science, and Google Scholar from inception to 12^th^ of March 2020. Cochrane’s Q and I-square statistics were used to assess the existence of heterogeneity between the included studies. We used the random-effects model to pool the odds ratios (ORs) at 95% confidence intervals (CIs).

**Results:** Seventeen articles out of 3009 citations were included. These contained 3189 patients, of whom 732 were severely affected (severe group) and 3189 were in non-severe group. Using the random-effects model, our meta-analyses showed that the odds of comorbidities, including COPD, DM, HTN, CVD, CKD, and symptoms, including dyspnea, dizziness, anorexia, and cough, were significantly higher among the severe group compared with the non-severe group. There were no significant changes in odds of CVA, liver disease, immunodeficiency/immunosuppression, fever, fatigue, myalgia, headache, diarrhea, sore throat, nasal congestion, sputum, nausea, vomiting, chest pain between the two groups.

**Conclusions:** Early recognition and intervention can be critical in management, and might stop progression to severe disease. Predictive symptoms and comorbidities can be used as a predictor in patients who are at risk of severe disease.

## Introduction

Coronaviruses have been responsible for emerging and re-emerging diseases through years, including severe acute respiratory syndrome (SARS) of 2003 and Middle-East respiratory syndrome (MERS) of 2012.^1,2^ Recently, a new coronavirus disease of 2019 (COVID-19) has been identified in Wuhan, China.^3^ Since then, it has spread to 210 countries/territories and infected nearly 2.5 million people around the world. The surge in the number of COVID-19 cases has caused an influx of patients to the hospitals. This has diminished health care resources, including personnel, hospital beds, and intensive care capacities.^4^ Therefore, it is essential to allocate resources for the patients who are at risk of severe disease.^5^

COVID_19 has become unpredictable due to the diversity of the symptoms and severity in different individuals. The clinical spectrum varies from asymptomatic disease to severe pneumonia and multi-organ failure and subsequent death.^6^ Some studies have investigated common clinical symptoms and the role of underlying diseases in the course of the disease. However, there is not a comprehensive opinion in this regard, and available studies are inconclusive. It is essential to gather all available data to provide more reliable information in this regard.

Here, we decided to conduct a comprehensive meta-analysis study to clarify predictive symptoms and the role of comorbidities in severely affected individuals.

## Methods

The Preferred Reporting Items for Systematic Reviews and Meta-analyses (PRISMA) was used in our systematic review and meta-analysis report.

### Search Strategy

We systematically searched online databases, including PubMed, Scopus, EMBASE, Web of Science, and Google Scholar from inception to 12^th^ of March 2020. The reference lists of previous reviews and eligible studies were manually checked to find additional studies that did not capture in online searches. The following both medical subject headings (MeSH) and keywords were used in the search strategy: “2019-nCoV disease” OR “2019 novel coronavirus disease” OR “COVID-19” OR “COVID19” OR “2019 novel coronavirus infection” OR “coronavirus disease 2019” OR “coronavirus disease-19” OR “2019-nCoV infection” OR “2019-nCoV” OR “2019 novel coronavirus” OR “2019 coronavirus” OR “novel coronavirus” OR (2019 AND coronavirus).

### Study selection

Two researches (M.A-K and N.Z) independently examined the relevant search results according to our inclusion criteria. Any discrepancies among them were resolved through consensus or discussion with a third researcher (RT). Our inclusion criteria included articles that were an original studies (cross-sectional, case-control, and cohort design); studies that investigated comorbidities such as chronic obstructive pulmonary disease (COPD), diabetes (DM), hypertension (HTN), cardiovascular disease (CVD), liver disease, chronic kidney disease (CKD), immunodeficiency/immunosuppression, cerebrovascular accident (CVA), symptoms including fever, cough, fatigue, myalgia, headache, diarrhea, sore throat, nasal congestion, sputum production, dyspnea, nausea, vomiting, anorexia, dizziness, chest pain. among others, final outcomes [includes discharge and death] in severe (as measured by disease severity criteria as severe/critical disease type or admitted to ICU or the use of mechanical ventilation) vs. non-severe (patients with mild or non-severe disease type) COVID-19 in English language; studies that reported sufficient data to extract number of intended clinical features for calculating odds ratios (ORs) with 95 % confidence intervals (95% CI) in patients with severe compared with non-severe group. Repeated articles, case reports, case series, literature reviews, letters, editorials, short communication, expert opinions, abstracts without full text, and coronavirus strains other than COVID-19 were excluded.

### Data Extraction and Quality Assessment

Two independent researchers (N.Z and M.A-K) extracted data from the selected studies for both narrative synthesis and statistical analysis. The following information were extracted: first author’s name, year of publication, study type, patient characteristics, study setting, number of investigated comorbidities, symptoms, final outcomes in both severe and non-severe groups. Discrepancies were resolved by consensus or discussion with a third author (P-N.S). To assess the quality of the included articles the Newcastle-Ottawa Scale (NOS) was used. This scale assessed the included studies on three aspects including participant selection, comparability, and exposure/outcome. We considered the quality assessment threshold with a NOS scored ≥ 7 being defined as good quality.

### Statistical Analysis

Random-effects meta-analysis was selected to estimate the pooled odds ratios (ORs) and its 95% confidence interval because of the diversity of patient’s definitions. Cochrane’s Q and I-square statistics were applied to assess heterogeneity across included studies. I^2^ above 70% and Cochrane’s Q test with P < 0.05 indicated the presence of significant heterogeneity. The potential evidence of publication bias was examined using Egger’s regression and Begg’s rank correlation tests and also was quantified using Duval & Tweedie’s trim and fill method. Series of sensitivity analyses (with leave-one-out method) were used to evaluate the robustness of our findings after excluding each included study on the pooled ORs of each feature. STATA version 12.0 (Stata Corp., College Station, TX) was used to conduct all statistical analyses.

## Results

As shown in Figure. 1, initial database searches yielded 3009 citations, after removing 1021 duplicate records, 1988 titles and abstracts were screened, and of which 249 full-text articles were retrieved. Finally, 17 articles met our inclusion criteria.^7-23^ Most of the selected articles were performed in China^7-22^, except for one that was conducted in Singapore.^23^ Number of patients in these studies was 3921 persons, of which 732 and 3189 fell in the severe and non-severe groups, respectively. The mean age of the patients in both groups ranged from 37 to 77 years. Most studies clearly stated that clinical data were initial assessments (on admission / before treatment). **Table 1** shows an overview of 17 included study characteristics in current systematic review.

**Table 1.**
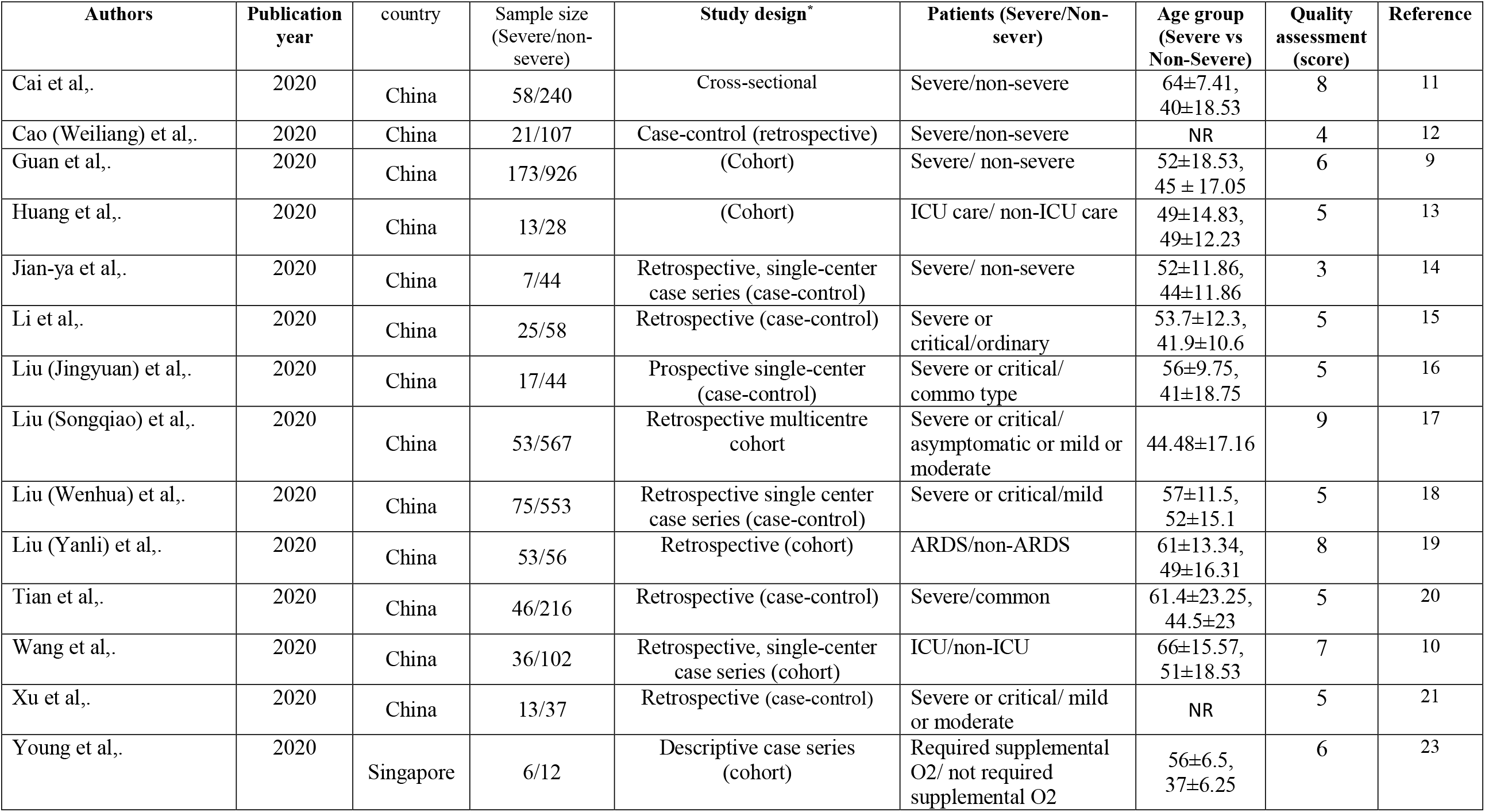
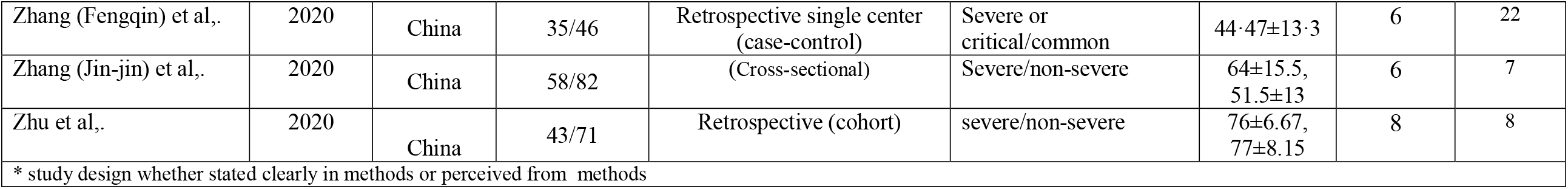
Characteristics of included studies

**Figure. 1.**
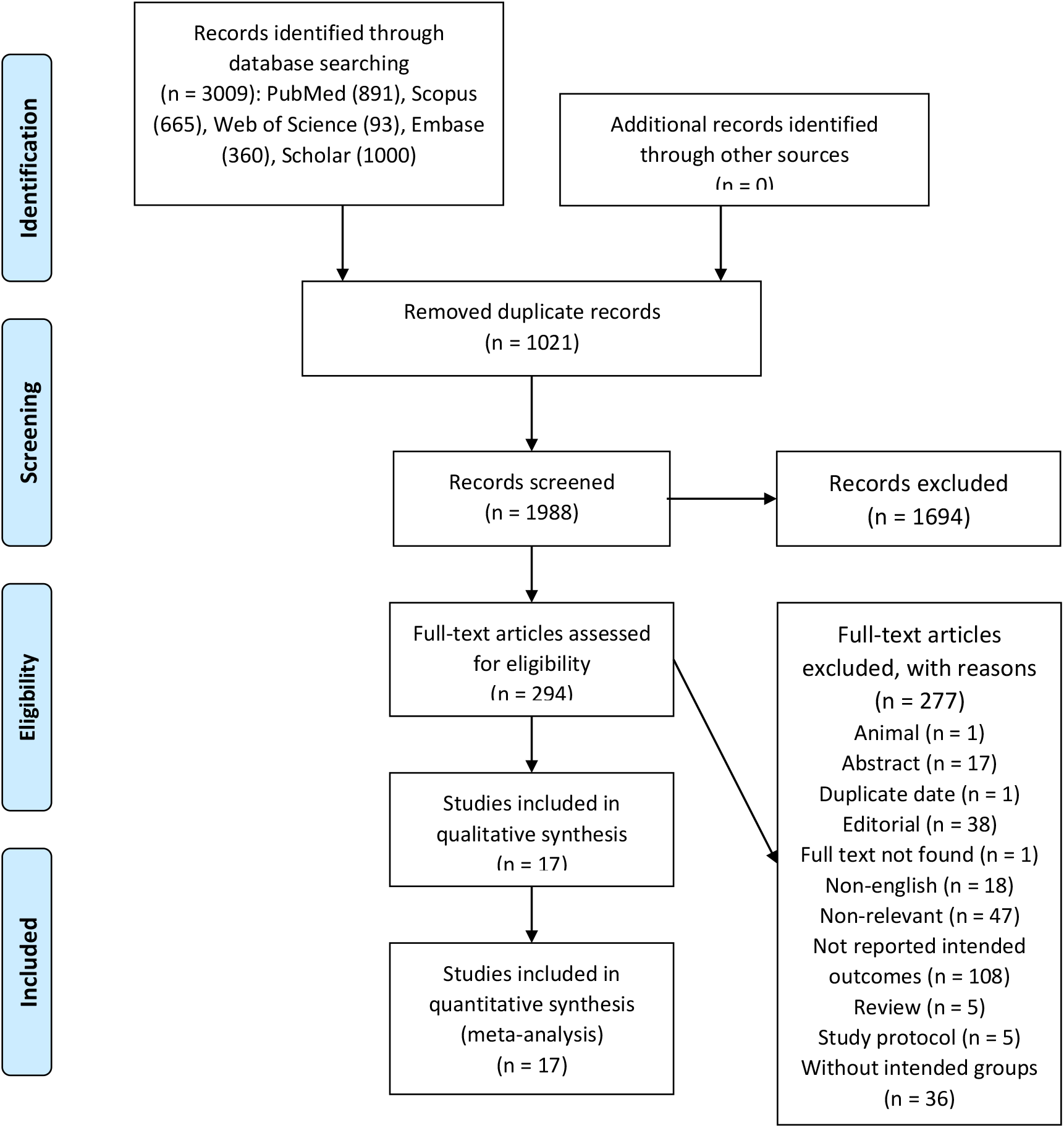
Study identificantion and selection process.

### Main outcomes

The pooled OR forest plots of comorbidities and clinical determinants in severe vs. non-severe patients with COVID-19 are illustrated in **Appendix 1: 2a-2d**.

### Comorbidities among severe vs. non-severe patients with COVID-2019

Using random-effects model, our meta-analyses showed that the odds of comorbidity condition presence such as CKD (n= 6, OR= 6.38, 95% CI 3.23–12.59, I^2^= 0.0%, Q-value= 4.52, P= 0.477), COPD (n= 8, OR= 4.07, 95% CI 2.30–7.22, I^2^= 0.0%, Q-value= 3.95, P= 0.786), DM (n= 9, OR= 3.54, 95% CI 1.79–7.01, I^2^= 58.4%, Q-value= 19.22, P= 0.014), CVD (n= 9, OR= 2.44, 95% CI 1.64–3.63, I^2^= 0.0%, Q-value= 6.43, P= 0.599), and HTN (n= 10, OR= 2.35, 95% CI 1.83–3.02, I^2^= 0.0%, Q-value= 7.38, P= 0.597) were significantly higher among severe group compared with the non-severe group.

No significant changes in odds of CVA (n= 4, OR= 3.94, 95% CI 0.88–17.59, I^2^= 53.1%, Q-value = 6.40, P= 0.094), liver disease (n= 6, OR= 1.25, 95% CI 0.35–4.41, I^2^= 40.2%, Q-value= 8.35, P= 0.138), immunodeficiency/immunosuppression, (n= 3, OR= 1.08, 95% CI 0.18–6.26, I^2^= 0.0%, Q-value= 0.38, P= 0.825) between the two groups were observed (**Appendix 1a, Fig: A-H**).

### Initial symptoms among severe vs. non-severe patients with COVID-2019

Our pooled results on symptoms indicated a significant higher odds of cough (n= 16, OR= 1.42, 95% CI 1.11–1.81, I^2^= 23.8%, Q-value= 19.68, P= 0.185), dyspnea (n= 14, OR= 5.94, 95% CI 3.37–10.45, I^2^= 66.2%, Q-value= 38.42, P< 0.001), anorexia (n= 4, OR= 3.18, 95% CI 1.46–6.93, I^2^= 9.4%, Q-value= 3.31, P= 0.346), and dizziness (n= 3, OR= 4.43, 95% CI 1.69–11.60, I^2^= 0.0%, Q-value= 0.40, P= 0.818) in the severe group compared with the non-severe group.

But, there were no differences in odds of fever (n= 16, OR= 1.20, 95% CI 0.94–1.52, I^2^= 4.6%, Q-value= 15.72, P= 0.401), fatigue (n= 11, OR= 1.49, 95% CI 0.99–2.24, I^2^= 57.4%, Q-value= 23.49, P= 0.009), myalgia (n= 8, OR= 1.21, 95% CI 0.87–1.69, I^2^= 0.0%, Q-value= 2.69, P= 0.912), headache (n= 9, OR= 1.01, 95% CI 0.70–1.46, I^2^= 0.0%, Q-value= 5.87, P= 0.662), diarrhea (n= 13, OR= 1.30, 95% CI 0.92–1.84, I^2^= 0.0%, Q-value= 9.54, P= 0.656), sore throat (n= 10, OR= 1.22, 95% CI 0.86–1.75, I^2^= 0.0%, Q-value= 8.50, P= 0.485), nasal congestion (n= 4, OR= 1.51, 95% CI 0.32–7.06, I^2^= 46.9%, Q-value= 5.65, P= 0.130), sputum production (n= 10, OR= 1.39, 95% CI 0.89–2.16, I^2^= 50.7%, Q-value= 18.24, P= 0.032), nausea (n= 7, OR= 1.13, 95% CI 0.58–2.19, I^2^= 30.9%, Q-value= 8.69, P= 0.192), vomiting (n= 5, OR= 1.92, 95% CI 0.61–6.00, I^2^= 15.9%, Q-value= 4.76, P= 0.313), and chest pain (n= 5, OR= 1.27, 95% CI 0.44–3.71, I^2^= 14.7%, Q-value= 4.69, P= 0.321) between the two groups (**Appendix 1b, Fig: A-P**).

### Final outcomes among severe vs. non-severe patients with COVID-2019

The pooled results indicated that odds of discharge were significantly higher among the non-severe group (n= 5, OR= 3.05, 95% CI 1.86–5.00, I^2^= 0.0%, Q-value= 1.90, P= 0.753) than the severe group. However, there were a significant increase in the odds of death (n= 6, OR= 10.56, 95% CI 3.09–36.09, I^2^= 66.6%, Q-value= 14.95, P= 0.011) in the severe group than the non-severe group, as we expected (**Appendix 1d, Fig: A and B**).

### Sensitivity analyses

We assessed the effect of each study on the robustness of the pooled ORs for each outcome by systematically removing each study from the meta-analyses. The findings of sensitivity analyses showed that there were no significant differences between the pre- and post-sensitivity pooled ORs for COPD, DM, HTN, CVD, liver disease, CKD, immunodeficiency/immunosuppression, fever, headache, chest pain, cough, myalgia, dyspnea, nausea, nasal congestion, discharge, and death. However after omitting Zhang (Jin-jin) et al.^7^, the study on CVA (OR= 6.47, 95%CI: 1.10, 38.08), Zhu et al.^8^, the study on sputum (OR=1.58, 95%CI: 1.03, 2.41), Guan et al.^9^, the study on sore throat (OR=1.73, 95%CI: 1.01, 2.98), Zhu et al.^8^, the study on fatigue (OR= 1.66, 95%CI: 1.10, 2.49), Zhang (Jin-jin) et al.^7^, the study on vomiting (OR= 3.64, 95%CI: 1.01, 13.07), Wang et al.^10^, the study on anorexia (OR=1.76, 95%CI: 0.52, 6.00), Wang et al.^10^, the study on dizziness (OR=2.92, 95%CI: -0.57, 14.88), Zhang (Jin-jin) et al.^7^, the study on diarrhea (OR=1.49, 95%CI: 1.01, 2.18), we found significant differences between pre- and post-sensitivity pooled ORs.

### Publication bias

The Egger’s regression and Begg’s rank correlation tests were conducted to find publication bias. These indicated no significant publication bias for COPD, HTN, CVD, liver disease, CKD, immunodeficiency/immunosuppression, CVA, fever, cough, myalgia, headache, diarrhea, sore throat, nasal congestion, sputum, nausea, vomiting, dizziness, chest pain, discharge, and death. Because there was evidence of potential publication bias on DM [Egger (P= 0.07), Begg (P< 0.01)], dyspnea [Egger (P= 0.02), Begg (P= 0.01)], fatigue [Egger (p= 0.04), Begg (P= 0.03)], anorexia [Egger (p< 0.01), Begg (P= 0.17)], We applied the non-parametric method (Duval and Tweedie) to estimate the results of censored studies. There were no significant changes between before and after including censored studies for DM, anorexia, dyspnea, and fatigue.

## Discussion

We identified comorbidities and initial clinical predictors for severe Covid-19 based on a meta-analysis of 17 studies. Our analysis identified symptoms, including cough, dyspnea, anorexia, and dizziness, and comorbidities, including COPD, DM, HTN, CVD, and CKD, as an indicator of severe disease.

The difference in initial clinical symptoms can be used as an indicator of the severe disease. The odds of dyspnea, dizziness, anorexia, and cough were 5.94, 4.43, 3.18, and 1.42 times, respectively, higher in severe patients than the non-severe group. Two previous meta-analyses reported different results in terms of the predictive symptoms. Jain et al. meta-analysis on seven studies reported dyspnea as the only predictor of severe COVID-19.^24^ However, Gong et al. found cough, fever, and dyspnea as the most clinical manifestation in severe cases.^25^

Guan et al. reported that about 25% of studied patients had at least one comorbidity.^26^ The impact of comorbidities might be explained by the tissue distribution of the SARS-Cov-2 receptor in host cells.^27,28^ ACE-2 is shown to be host receptor for the binding of SARS-Cov-2 virus to the cell, similar to SARS-CoV.^29^ COPD was higher among severe patients in our study, which is consistent with previous studies.^24,26^ This impact might be due to the higher expression of the ACE-2 receptor among COPD patients.^30,31^ CKD was related to severe disease in our study, as well. *Cheng et al*. reported higher in-hospital acute kidney injury among patients with abnormal baseline creatinine, which is a determinant of poor prognosis.^32^ Also, there is a higher probability for CKD patients to become critically ill in the course of COVID-19, which might be due to consuming ACE inhibitors and ARBs in this group. *Diaz et al*. discussed the potential increase of ACE 2 receptors in the patients consuming ACE inhibitors and ARBs, as shown in experimental animals.^33^ DM, HTN, CVD can be a determinant of the severe status of the diseases, in concordance with previous studies as reported by *Li et al*.^34^ due to the increase in ACE-2 receptors in these populations.^35^

Investigated complications were higher among the severe cases.

The importance of such studies is the early recognition of high risk and critically ill patients. By identifying influential comorbidities on disease severity, we can recognize vulnerable population earlier and provide targeted care.

Comorbidities and predictive symptoms can be applied in clinical practice. Comorbidities can be a good predictor of severe disease. It can be used in recognizing more vulnerable population, and to prevent them from getting the disease by emphasizing protective measures and education.

Predictive symptoms and comorbidities can be used to recognize patients who are at risk of severe disease. Sun et al. mentioned that early and aggressive intervention is an effective way to reduce mortality.^36^ Early recognition and intervention can be critical in management, and might stop progression to severe disease. Therefore, logical allocation of available resources can be very important at this stage.

The present study had some limitations that should be acknowledged. First, definitions of severity across the included studies were inconsistent. However, overall results showed that the odds of death were 10.56 times higher among severe than the non-severe group. This means that the chance of death was significantly higher in the severe group, which implies that patients were in a more critical situation in severe than the non-severe group. second, our analysis was based on small number of studies in some outcomes, however, the relevant results should be interpreted with caution. Third, most of the included studies were from China and may have population biases. Fourth, Heterogeneity across included studies on some outcomes was another limitation and sensitivity analysis was done to address this limitation.

## Data Availability

data regarding the manuscript will be available as request

